# Omicron Spread in Vaccinated Jurisdictions: a Statistical Study

**DOI:** 10.1101/2022.03.15.22272430

**Authors:** Serge Dolgikh

## Abstract

A correlation hypothesis between the level of vaccination and the rate of spread of the new Covid-19 variant is investigated based on the case and vaccination data from European and North American jurisdictions available in the public domain at the time point of past the crest of the Omicron wave in most jurisdictions. Statistical variables describing the rate of the spread based on observed new case statistics defined and discussed. An unexpected moderate positive correlation between the rate of the variant spread measured by two related parameters and vaccination level based on the dataset in the study is reported. While negative correlation was not statistically excluded, the analysis of the data in the study statistically excluded moderate to strong negative correlation. The results of this work, if confirmed by further independent studies can have implications for development of policies aimed at controlling future course of the Covid-19 pandemic.

## 1 Introduction

A novel variant of Covid-19, branded Omicron was detected in November, 2021 [1,2] spreading rapidly across the globe. In this work we set out to examine the question, how do the rate of spread that is reflected in the observed epidemiological variables across national and subnational jurisdictions relate to public health measures such as the rate of vaccination.

To reduce the influence of multiple factors, jurisdictions with comparable socio-economical parameters were chosen, comprising a set of national and subnational public health jurisdictions in Europe and North America.

The data for the analysis was obtained from the open to the public sources [3-5].

## 2 Data

We examined a correlation hypothesis between the vaccination level in a number of European and North American jurisdictions (parameter *vacc*) and the rate of spread of the new Covid-19 variant, Omicron.

The rate of spread was measured by two variables: the rate of spread (variable *rate_spr*), defined as the ratio of new case counts, the peak count of the Covid-19 new cases curve (*case-peak*) to the preceding trough of the curve (*case-min*). For the case counts, seven days average was used in most cases. The variable defines an invariant factor that is not dependent on a specific choice of a calibrating value of the case number in the jurisdictions, and is determined only by the factors in the jurisdiction.

The second variable, *rate-max* was defined as the peak number of cases (*case-peak*) per 1 million capita.

The spread rate variables could not be considered as fully independent as both used the same factor, *case-peak*. Yet using both variables can improve the confidence in the finding as they reflected different aspects of the new case statistics, one being dimensionless and fully controlled by the jurisdictional factors; whereas the second parameter, *rate-max* contained a common calibrating factor (capita count) across multiple jurisdictions.

Vaccination level per jurisdiction was reported as the number of administered doses per capita of population. The number included administered single doses and cannot be considered as an exact measure of the fully vaccinated eligible population in the jurisdiction.

For the correlation analysis, we selected a set of 30 European and North American jurisdictions as: 1) European jurisdictions [1] by the level of vaccination at the time of the analysis, and two North American jurisdictions (Ontario, Quebec, Canada). Jurisdictions with population less than one million were excluded from the analysis.

Vaccination and case statistics were obtained from publicly available sources [3-6]. The data on the national case statistics (*case-peak, case-min*) was recorded at two time points:

### Timepoint 1 (Interim)

17-18.12.2021. At this time point some or many jurisdictions in the dataset were still going through the Omicron wave and the analysis can be considered as interim.

### Timepoint 2 (Past crest)

22-24.01.2022. At this time point most jurisdictions have reached the peak of the Omicron wave or were past it, and the dataset for a statistical correlation analysis can be compiled in the final form.

The resulting dataset of at the first (Interim) timepoint is given in Table 1. The data is available upon request.

**Table 1.**
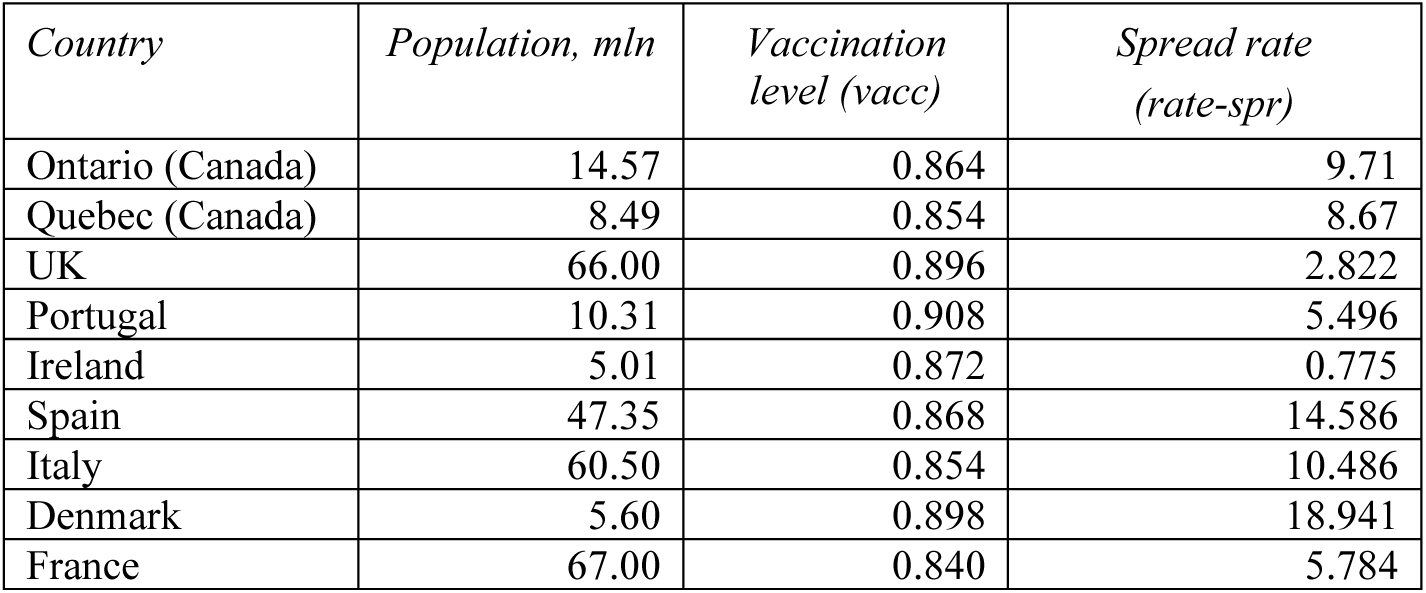
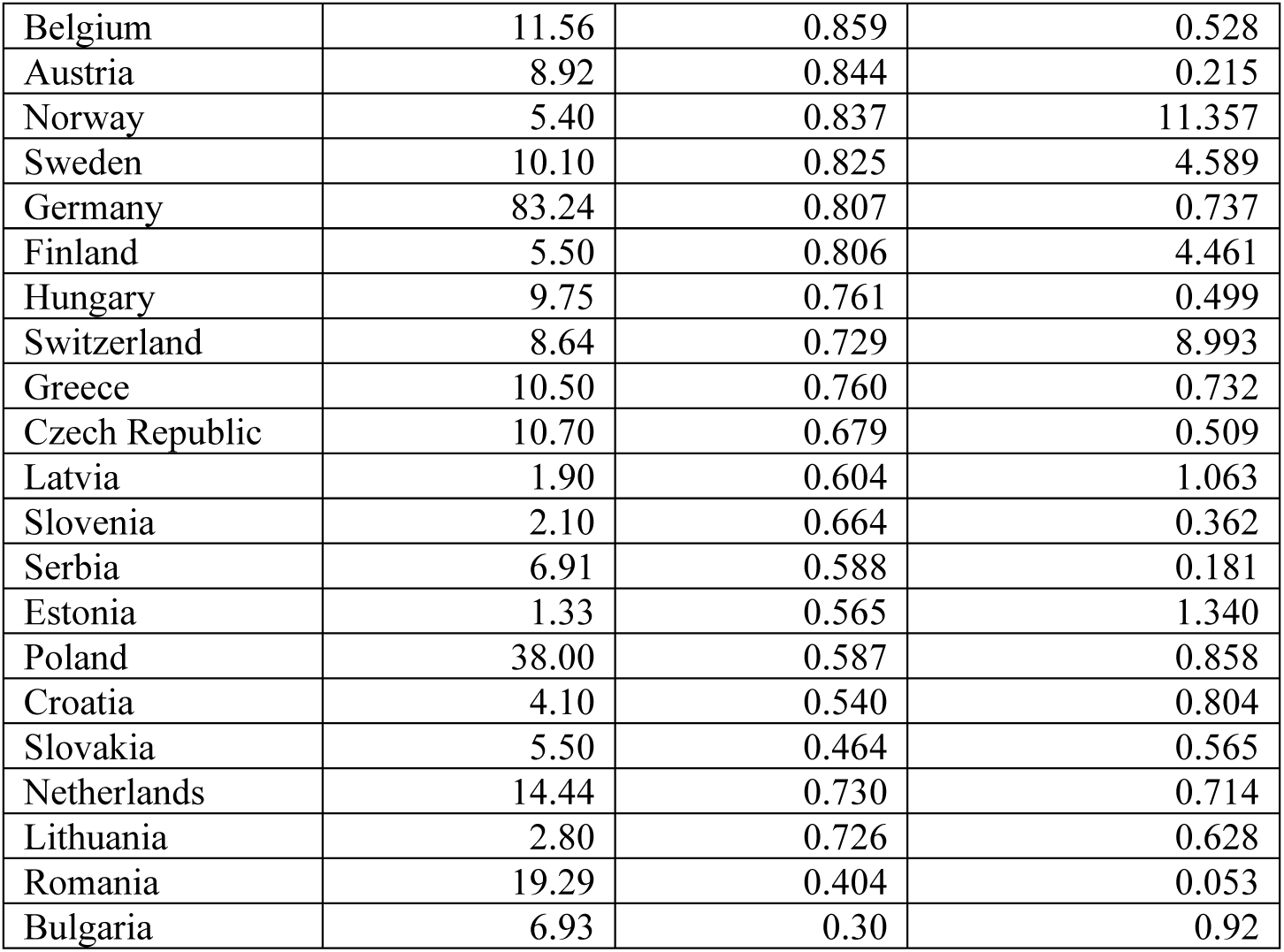
National Covid-19 vaccination and spread rate statistics (Interim timepoint)

| *Country* | *Population, mln* | *Vaccination level (vacc)* | *Spread rate (rate-spr)* |
| --- | --- | --- | --- |
| Ontario (Canada) | 14.57 | 0.864 | 9.71 |
| Quebec (Canada) | 8.49 | 0.854 | 8.67 |
| UK | 66.00 | 0.896 | 2.822 |
| Portugal | 10.31 | 0.908 | 5.496 |
| Ireland | 5.01 | 0.872 | 0.775 |
| Spain | 47.35 | 0.868 | 14.586 |
| Italy | 60.50 | 0.854 | 10.486 |
| Denmark | 5.60 | 0.898 | 18.941 |
| France | 67.00 | 0.840 | 5.784 |
| Belgium | 11.56 | 0.859 | 0.528 |
| Austria | 8.92 | 0.844 | 0.215 |
| Norway | 5.40 | 0.837 | 11.357 |
| Sweden | 10.10 | 0.825 | 4.589 |
| Germany | 83.24 | 0.807 | 0.737 |
| Finland | 5.50 | 0.806 | 4.461 |
| Hungary | 9.75 | 0.761 | 0.499 |
| Switzerland | 8.64 | 0.729 | 8.993 |
| Greece | 10.50 | 0.760 | 0.732 |
| Czech Republic | 10.70 | 0.679 | 0.509 |
| Latvia | 1.90 | 0.604 | 1.063 |
| Slovenia | 2.10 | 0.664 | 0.362 |
| Serbia | 6.91 | 0.588 | 0.181 |
| Estonia | 1.33 | 0.565 | 1.340 |
| Poland | 38.00 | 0.587 | 0.858 |
| Croatia | 4.10 | 0.540 | 0.804 |
| Slovakia | 5.50 | 0.464 | 0.565 |
| Netherlands | 14.44 | 0.730 | 0.714 |
| Lithuania | 2.80 | 0.726 | 0.628 |
| Romania | 19.29 | 0.404 | 0.053 |
| Bulgaria | 6.93 | 0.30 | 0.92 |

## 3 Results

### 3.1 Interim Analysis: Statistically Significant Positive Correlation

Pearson’s correlation coefficient calculated for variables *vacc, rate-spr* as recorded in Table 1 produced a moderate positive value of **0.531**, statistically significant with P-value of 0.0040, 95% CI [0.204, 0.751], excluding the null hypothesis, correlation value of 0, with a 95% confidence.

A positive value of the correlation coefficient means that higher level of vaccination is correlated with higher rate of spread in the jurisdiction.

This result is confirmed by a direct observation of the case curves in Fig.1,2. Type 1 case curve, rapid acceleration type (Fig.1) was observed for 7 jurisdictions in the highest vaccination rate group out of ten (except: Ireland, Belgium, Austria).

**Fig.1.**
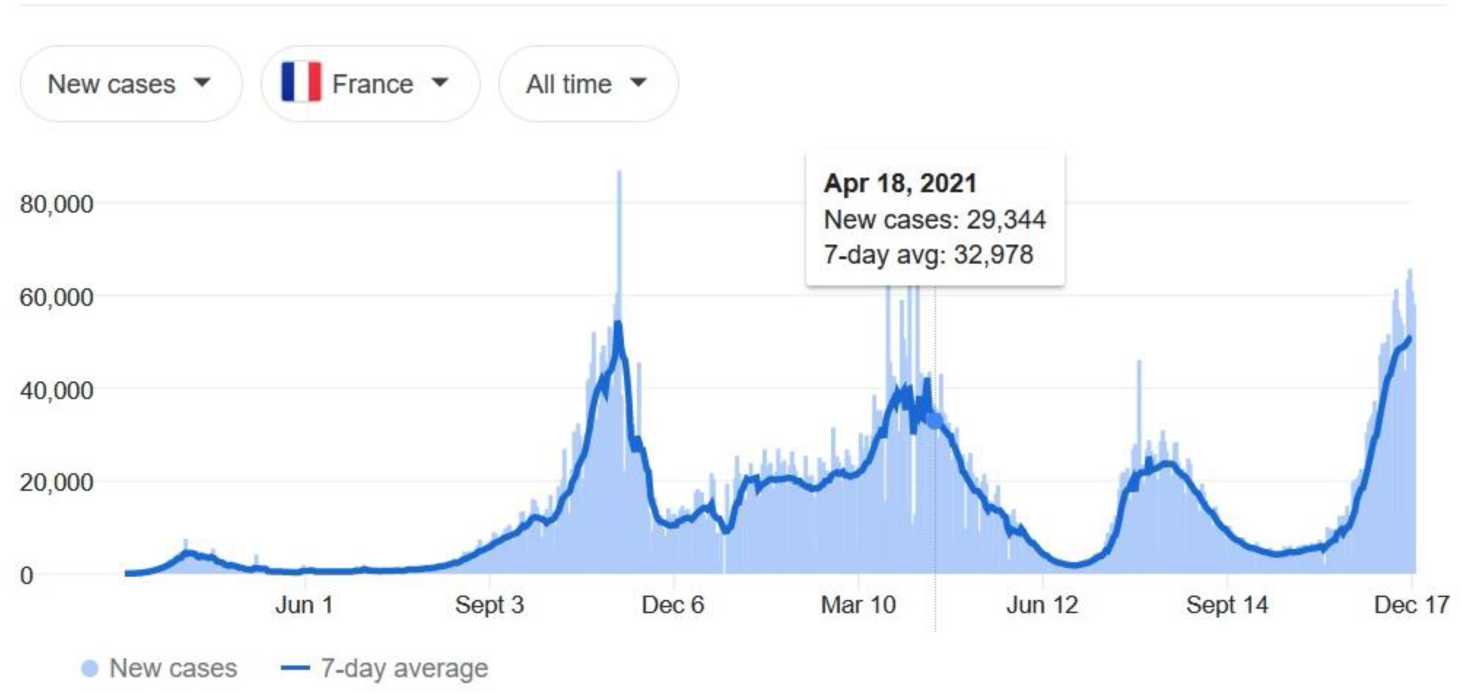
Type 1 Case curve (France)

**Fig.2.**
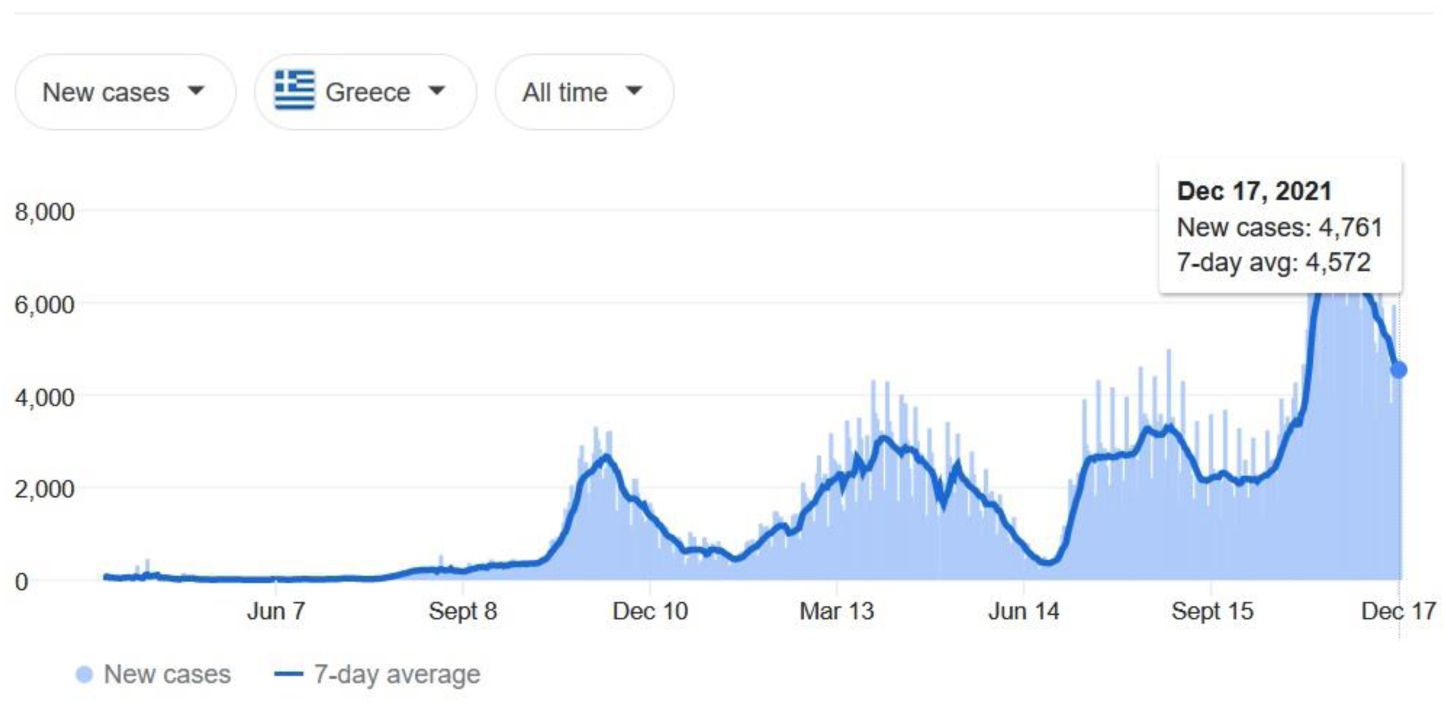
Type 2 Case curve (Greece)

Type 2 curve (descending type, Fig.2) was observed for 8 out of 10 lower vaccination rate group jurisdictions, with the cases of Latvia and Estonia, inconclusive.

#### Caveats

1. It is possible that some jurisdictions are not yet experiencing an accelerating wave of Omicron (i.e., a delayed epidemical cycle, or an inter-cycle interval, between Delta and Omicron waves). A more detailed analysis of cases by variant type would be useful to identify such scenarios.
2. It is possible that some cases are reported less frequently in these jurisdictions for a number of reasons.

To address these concerns and improve the confidence in the findings on the correlation hypothesis, it can be recommended to update the dataset and repeat the analysis at a future timepoint when most or all jurisdictions in the dataset have passed the local crest of an infectious wave.

### 3.2 Past-crest Analysis: Statistically Significant Exclusion of a Significant Negative Correlation

The analysis at the interim timepoint in the preceding section was repeated at another time point, when all jurisdictions in the set passed the peak of the omicron wave. At that time case data for the ascending arc of the epidemic cycle was available for all jurisdictions that allowed to perform the correlation study with a complete set of statistics for both spread variables, *rate-spr* and *rate-max*.

As with the interim case, a positive value of the correlation coefficient was recorded for both spread variables, *rate-spr, rate-max*: 0.276, 0.243 respectively. The p-value of the *rate-spr* correlation was 0.14, excluding negative correlation with a confidence of over 85%.

The value of correlation coefficient obtained with the full curve, past the crest of the cycle statistics was significantly lower than in the interim case. This is understandable as at the time of interim analysis many of the local epidemiological cycles have not yet reached local peaks and the statistics could be expected to be skewed, as commented earlier.

While the confidence of a positive correlation in the analysis has not reached the commonly accepted level of confidence, it was noticeably high (0.86). Moreover, the 95% confidence interval of the correlation coefficient for *rate-spr* variable was: [– 0.094; 0.579] i.e., confidently excluding a negative correlation with an absolute value greater than 0.1.

## 4 Conclusions

In this study we looked into a relation between variables characterizing the spread of an infection in health jurisdictions and certain parameters of public health policy, such as mass vaccination level. A novel, invariant parameter determined entirely by the factors specific to the jurisdiction describing the rate of spread proposed and a statistical approach to an analysis of the correlation demonstrated.

The consistency of findings of the statistical analysis between the interim and peak timepoints though interesting, may not have strong significance; depending on the choice of an interim point, epidemiological scenarios developing along different temporal cycles in different jurisdictions can produce different results. The consistency therefore may only be a matter of a coincidence in the choice of an interim timepoint.

On the other hand, the consistency of the results obtained with two variables describing the rate of infectious spread, *rate-spr* and *rate-max* improves the confidence in the findings. While not entirely independent, they describe propagation of the infection in the population of the jurisdictions in the dataset from different angles, and obtaining consistent results improves overall confidence.

Let us consider the options logically possible for the correlation hypothesis: a) a pronounced, i.e., moderate or strong negative correlation; b) an uncertain correlation, in a range of values of the correlation coefficient [– α, α] where α < 1: a small positive value, and c) a pronounced positive correlation. Then, the first possibility appears to be excluded by the results of the study by the constraints on the CI interval, Section 3.2.

This conclusion leaves two possibilities: an indifferent rate of spread with respect to the vaccination level, option b) above; and a positive correlation that can be mild, moderate or strong, option c). The second possibility is not excluded by these results, with the values of correlation coefficient allowed in the interval [–0.094, α], however according to the constraints on CI it is not preferred. The preferred range of values falls into the center of the confidence interval, approximately 0.3 that is, a moderate positive correlation.

Either of these possibilities can be seen as somewhat counter to the expectation (note that the analysis here deals with the factors of spread rather than the impacts of the epidemics on the population, such as morbidity and others). Both, if confirmed with an improved confidence by further independent studies, could have implications for development of public health policies. However, in the authors view it is the possibility and statistical preference for the last variant, c) that should merit an immediate attention of the research community at this time.

If confirmed by a more representative study, it may point at a possibility of an accelerated production of vaccine-resistant variants, that can propagate at an accelerated rate in highly immunized populations. Such a possibility was pointed out in some scenarios of interaction of a rapidly mutating infectious agents with a mass vaccinated population [7]. Possible mechanisms of production of resistant strains were discussed in a number of studies [8-10].

For this reason, in the authors view, the topic in this study and the methods proposed to approach it merit further attention of the research community.

## Data Availability

All data produced in the present study are available upon reasonable request to the authors

https://www.statista.com/statistics/1196071/covid-19-vaccination-rate-in-europe-by-country

https://www.google.com/covid19-map/

https://covid19tracker.ca/vaccinationtracker.html

